# ECG predictors of AF: a systematic review (Predicting AF in Ischaemic Stroke-PrAFIS)

**DOI:** 10.1101/2023.05.21.23290310

**Authors:** Alexander Berry-Noronha, Luke Bonavia, Edmund Song, Daniel Grose, Damian Johnson, Erin Maylin, Ernesto Oqueli, Ramesh Sahathevan

**Author notes:** Corresponding Author: Alexander Berry-Noronha. Disclosures: none. Funding: none.

## Abstract

In 25% of patients presenting with embolic stroke, a cause is not determined. Atrial fibrillation (AF) is a commonly identified mechanism of stroke in this population, particularly in older patients. Conventional investigations are used to detect AF, but can we predict AF in this population and generally? We performed a systematic review to identify potential predictors of AF on 12-lead electrocardiogram (ECG).

**Method:** We conducted a search of EMBASE and Medline databases for prospective and retrospective cohorts, meta-analyses or case-control studies of ECG abnormalities in sinus rhythm predicting subsequent atrial fibrillation. We assessed quality of studies based on the Newcastle-Ottawa scale and data were extracted according to PRISMA guidelines.

**Results:** We identified 42 studies based on our criteria. ECG patterns that predicted the risk of developing AF included interatrial block, P-wave terminal force lead V1, P-wave dispersion, abnormal P-wave-axis, abnormal P-wave amplitude, prolonged PR interval, left ventricular hypertrophy, QT prolongation, ST-T segment abnormalities and atrial premature beats. Furthermore, we identified that factors such as increased age, high CHADS-VASC, chronic renal disease further increase the positive-predictive value of some of these parameters. Several of these have been successfully incorporated into clinical scoring systems to predict AF.

**Conclusion:** There are several ECG abnormalities that can predict AF both independently, and with improved predictive value when combined with clinical risk factors, and if incorporated into clinical risk scores. Improved and validated predictive models could streamline selection of patients for cardiac monitoring and initiation of oral anticoagulants.

## Introduction

Stroke is a leading cause of long-term disability and mortality. There is substantial impact on quality of life, burden of care, and significant ongoing financial burden for healthcare systems. Proportionally, 75-89% of strokes occur at ages 65 or older (1). Older people suffer more severe strokes, are more likely to require institutional care and experience higher mortality. The risk of stroke secondary to atrial fibrillation (AF) is highest in this group increasing from 1.5% below 60, to 23.5% above 80 years (2).

Atrial fibrillation (AF) was first recognised 200 years ago and is an established stroke risk factor (2). It is the most common sustained cardiac rhythm disorder in the world with more than 30 million people affected globally (3). Approximately 25% of ischaemic strokes have been attributed to AF (4) with an associated increase in morbidity and mortality. AF-related stroke is likely associated with atrial cardiomyopathy, leading to stasis and clot formation in the left atrial appendage (5). It has been suggested that atrial hypomobility and impaired atrial endothelial function contribute to stroke occurrence in the absence of demonstrable AF(5). Subsequently, AF may represent a marker of atrial cardiomyopathy. The temporal association between AF episodes and stroke is however undetermined. Studies of patients with implantable cardiac devices suggests a delay between clot formation and subsequent stroke (6).

Approximately 25% of strokes have no clear cause at presentation and are referred to as ‘cryptogenic strokes ‘(7). In 2014, the term “embolic stroke of undetermined source “ (ESUS) was coined. ESUS denotes cortical strokes that are thought to be secondary to an embolus, but with no identifiable proximal source of a clot. Studies based on implantable loop recorders in ESUS patients demonstrated AF in at least 25% (8). Implantable cardiac monitoring devices may identify AF, but routine use is resource intensive and invasive(9).

Oral anticoagulation (OAC) is standard of care in preventing stroke in AF (10). There is no evidence to support empiric anticoagulation over aspirin in the ESUS population. Two large studies failed to demonstrate a reduction in stroke recurrence with either dabigatran or rivaroxaban (11,12). In one study, the lower stroke recurrence rate of 6.6% in the dabigatran group vs 7.7% in the aspirin group (HR=0.85; 95% CI 0.69-1.03, p=0.10) was not statistically significant and offset by increased bleeding.

Predictive risk scores and machine learning algorithms have been used to predict AF risk. Predictive scoring systems such as the Framingham heart study (FHS) and Cohorts for Ageing Research in Genomic Epidemiology model for AF (CHARGE-AF) incorporate clinical, electrocardiography (ECG) and transthoracic echocardiogram (TTE) criteria. These predictive systems have demonstrated poor clinical utility suggesting that better predictive scoring systems need to be developed to identify patients at risk of paroxysmal AF(13). Improved identification of the population of interest may help identify patients who benefit from streamlined long-term cardiac monitoring or empirical anticoagulation.

We conducted a systematic review of ECG markers and associated risk of developing AF. These ECG patterns could be used in developing more accurate AF-predictive systems. Subsequently, this could lead to focused use of loop-recorders or informed empirical use of OAC in patients with ESUS.

## Methodology

### Inclusion criteria

We included prospective, retrospective and case-control cohort studies published from 2014 until August 2021. We included cohorts who presented from the community (e.g. to primary care facilities or from pre-employment screening) in addition to those presenting with stroke. These cohorts were felt to best represent our population of interest, We restricted studies to those where the exposure of interest included a 12-lead ECG with the primary outcome being incidence of new AF, and where exposure was measured as odds ratio (OR), hazard ratio (HR) or relative risk (RR).

### Exclusion criteria

We excluded studies which included patients with known AF. We also excluded cohorts with pre-existing cardiac disease as this was not felt to be generalisable to our population of interest. We excluded editorials, review articles or correspondence/letters to the editor. We also excluded studies which relied on exposures other than a 12-lead ECG (e.g. Holter monitor).

### Search methodology

Our search was conducted on EMBASE and Medline using key search terms consisting of ‘electrocardiography ‘, ‘atrial fibrillation ‘, ‘risk factor ‘, ‘predictive value ‘ (see Appendix 1 for full search terminology). Further studies were identified from the references of selected articles. We limited our search to articles published in English. Following an initial search, abstracts were screened to determine suitability for inclusion. This was independently validated by a second reviewer.

The information from these studies was amalgamated, and where the same predictors were analysed, comparisons were made. Key data that were screened included study type, ECG abnormality and method of measuring this, number of participants, follow-up duration, quantified risk of AF and potential biases. This information was collected in accordance with the guidelines for meta-analyses and systematic reviews of observational studies for statement and the Preferred Reporting Items for Systematic Reviews and Meta-Analyses (PRISMA) for screening studies.

### Selection process

Quality assessment was based on the Newcastle Ottawa quality assessment scale according to (1) selection of patients, (2) comparability of groups or cohorts, and (3) exposure evaluation for case control studies and outcome evaluation for cohort studies.

### Data collection process

Data were entered in pre-designed spreadsheets using Microsoft Excel. Abstracts were retrieved as complete manuscripts and assessed against the inclusion criteria. Data extracted included publication details (author, year), study design, follow-up duration, endpoints, quality score, characteristics of the population and sample size. Two reviewers (AB and LB) independently reviewed the data, with disagreements resolved with input from a third reviewer (RS). Principally, information was extracted according to PRISMA guidelines and the PRISMA 2020 checklist was used. Data was then arranged by ECG pattern.

## Results

We identified 1743 individual articles. Based on application of inclusion and exclusion criteria we identified 45 articles that were analysed. The complete PRISMA spreadsheet detailing our search results is shown in Figure 1.

**Figure 1.**
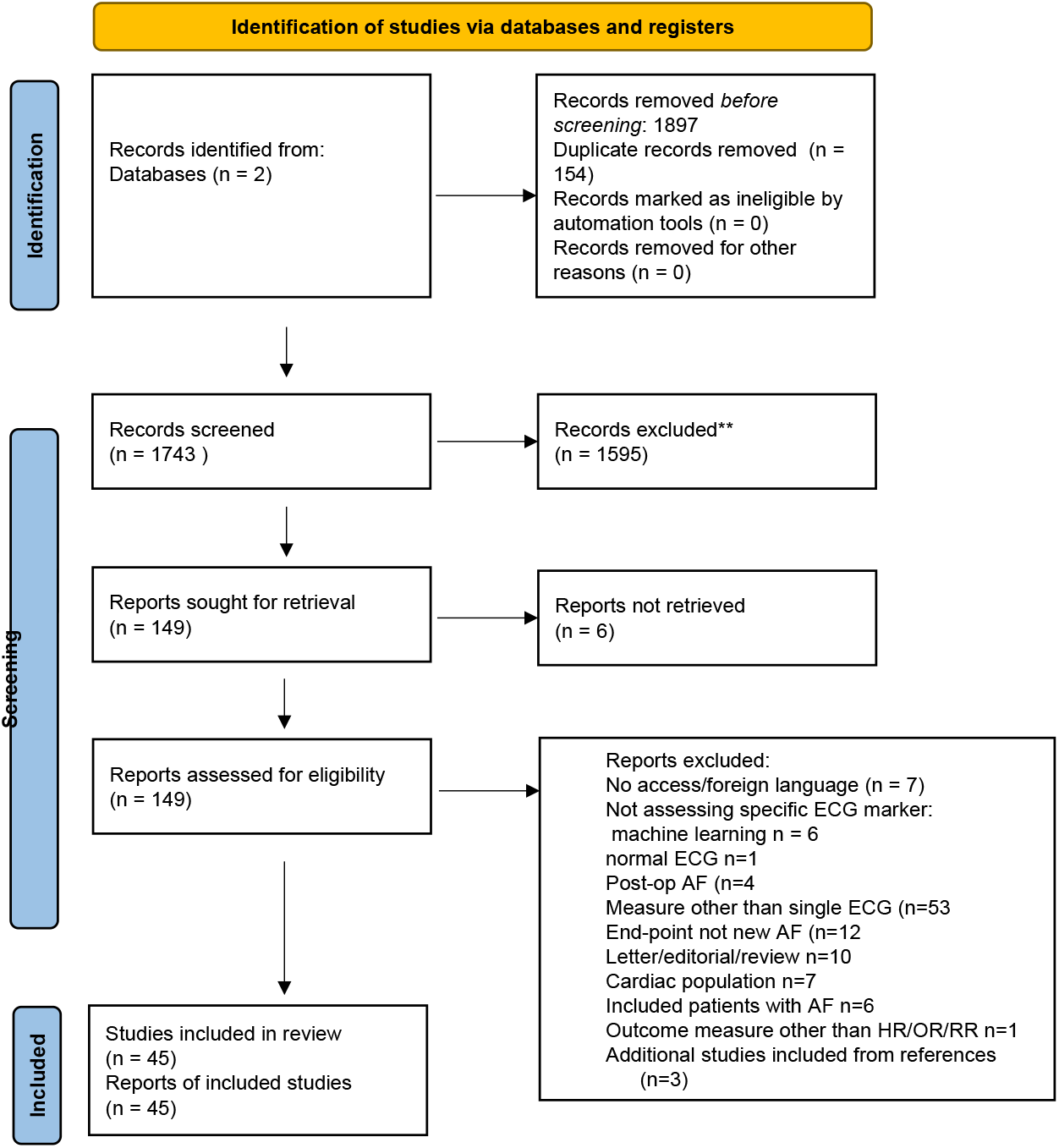
Systematic review search results

Our results are presented based on the type of ECG pattern and abnormality. Figure 2 provides an explanation of ECG patterns. All studies included in this review are tabulated in Table 1.

**Table 1.**
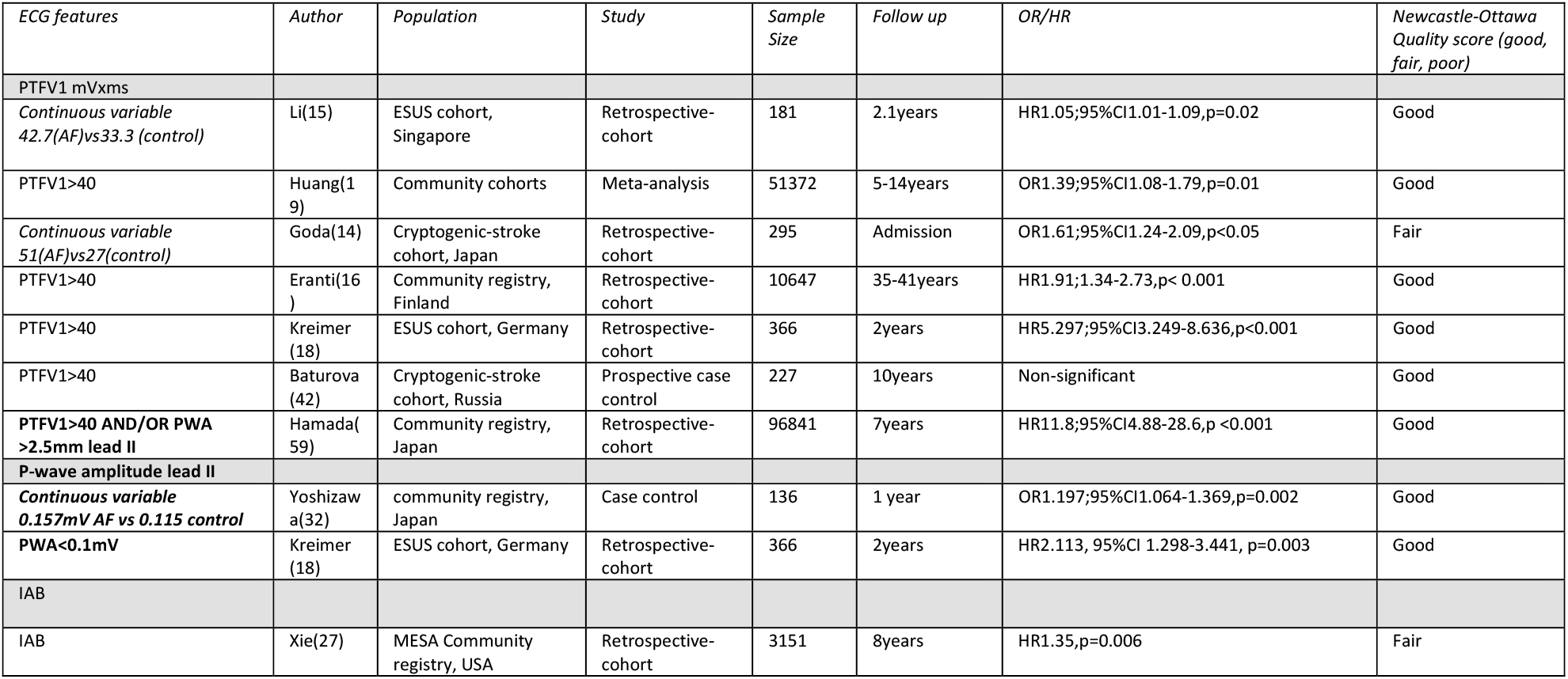
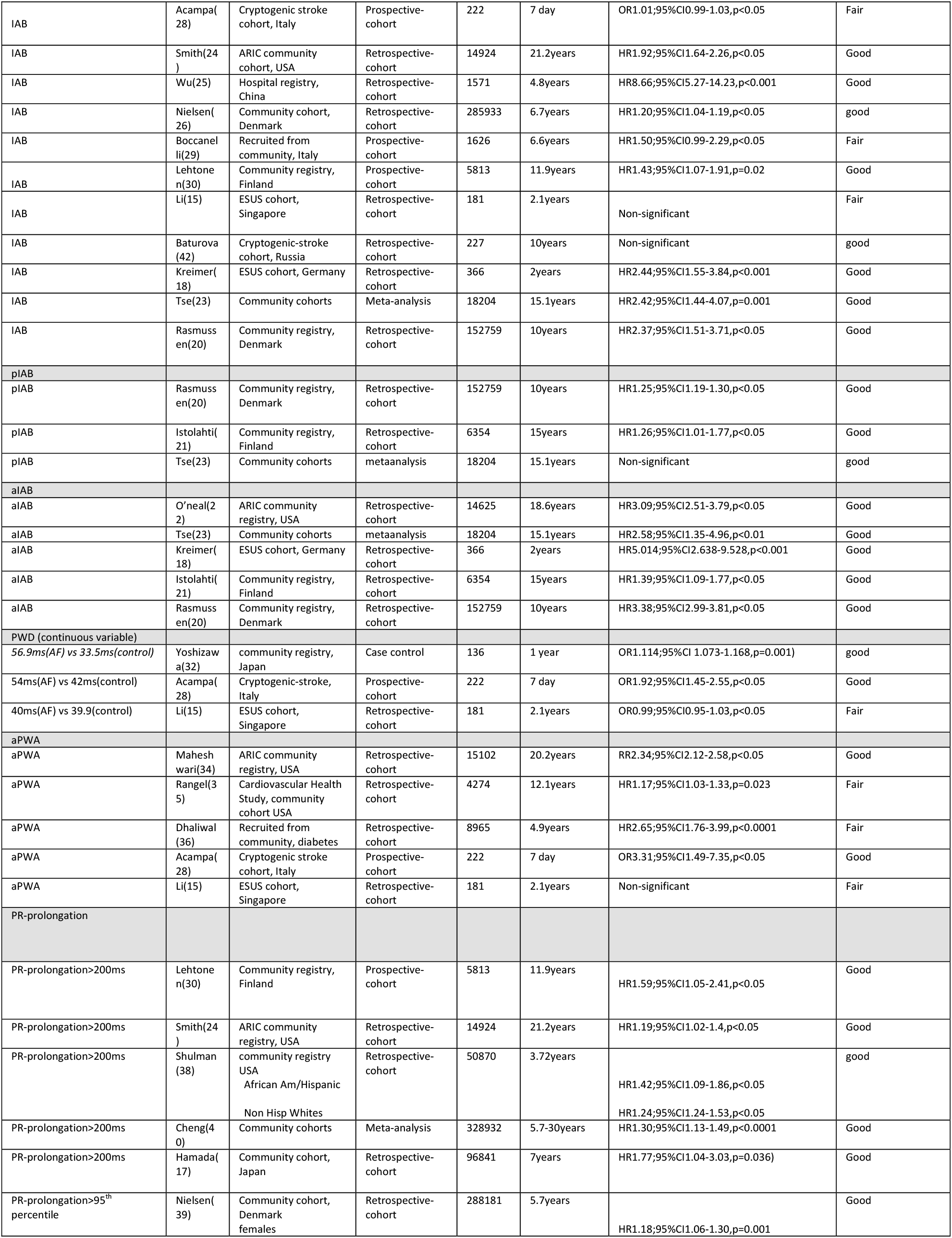
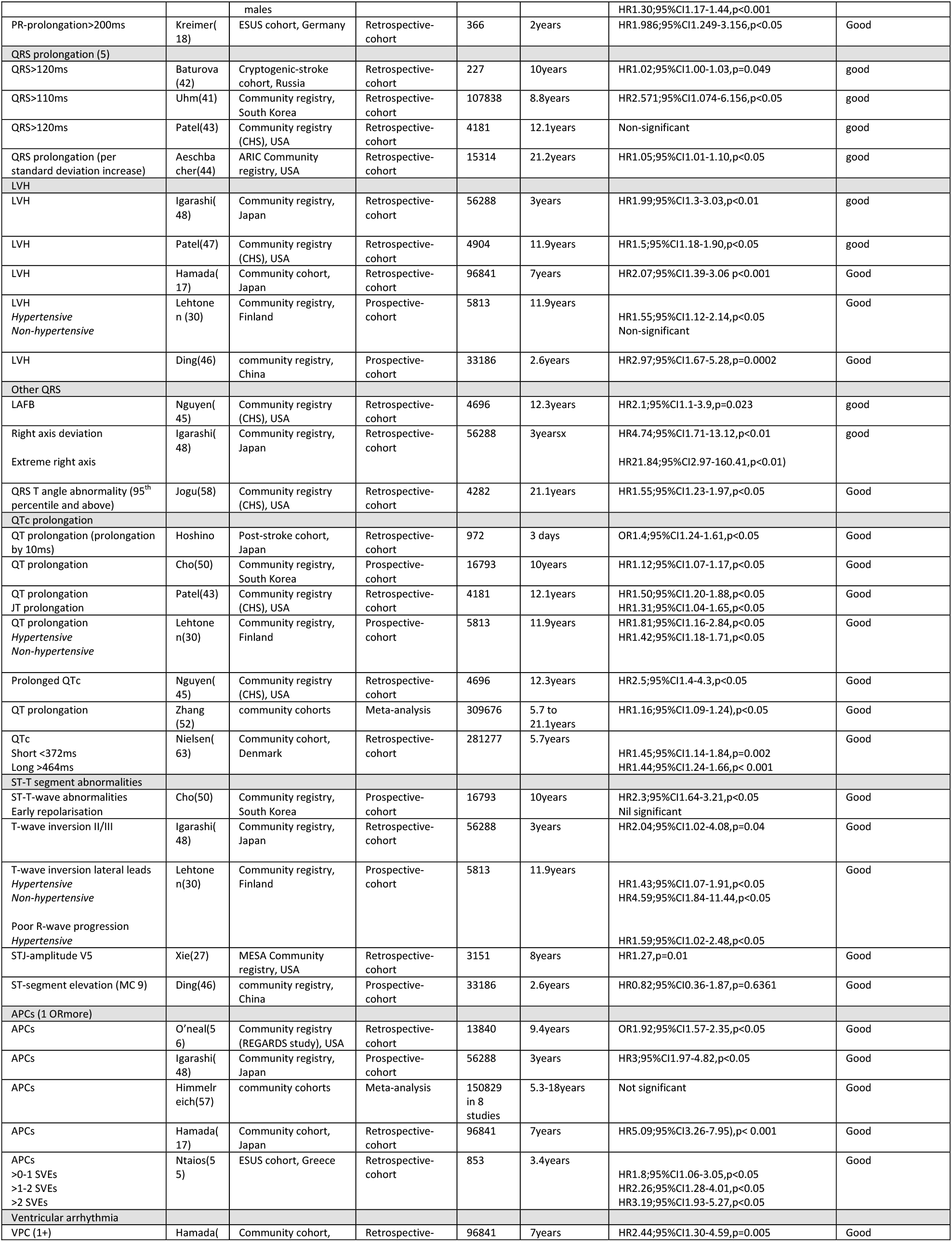
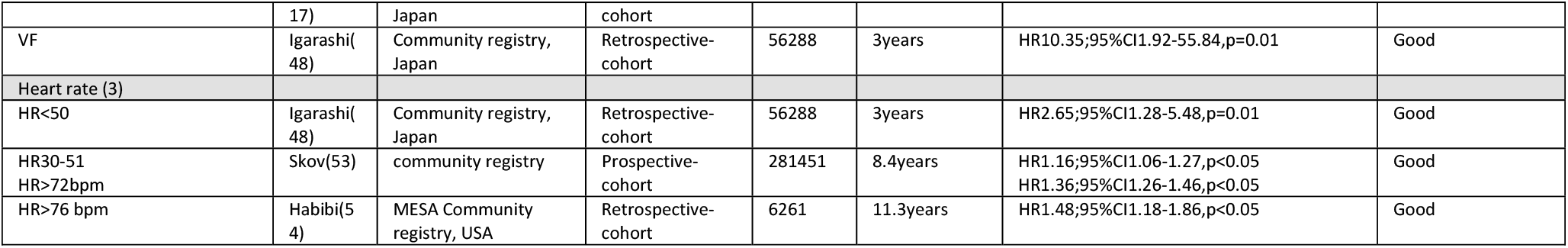

**Figure 2.**
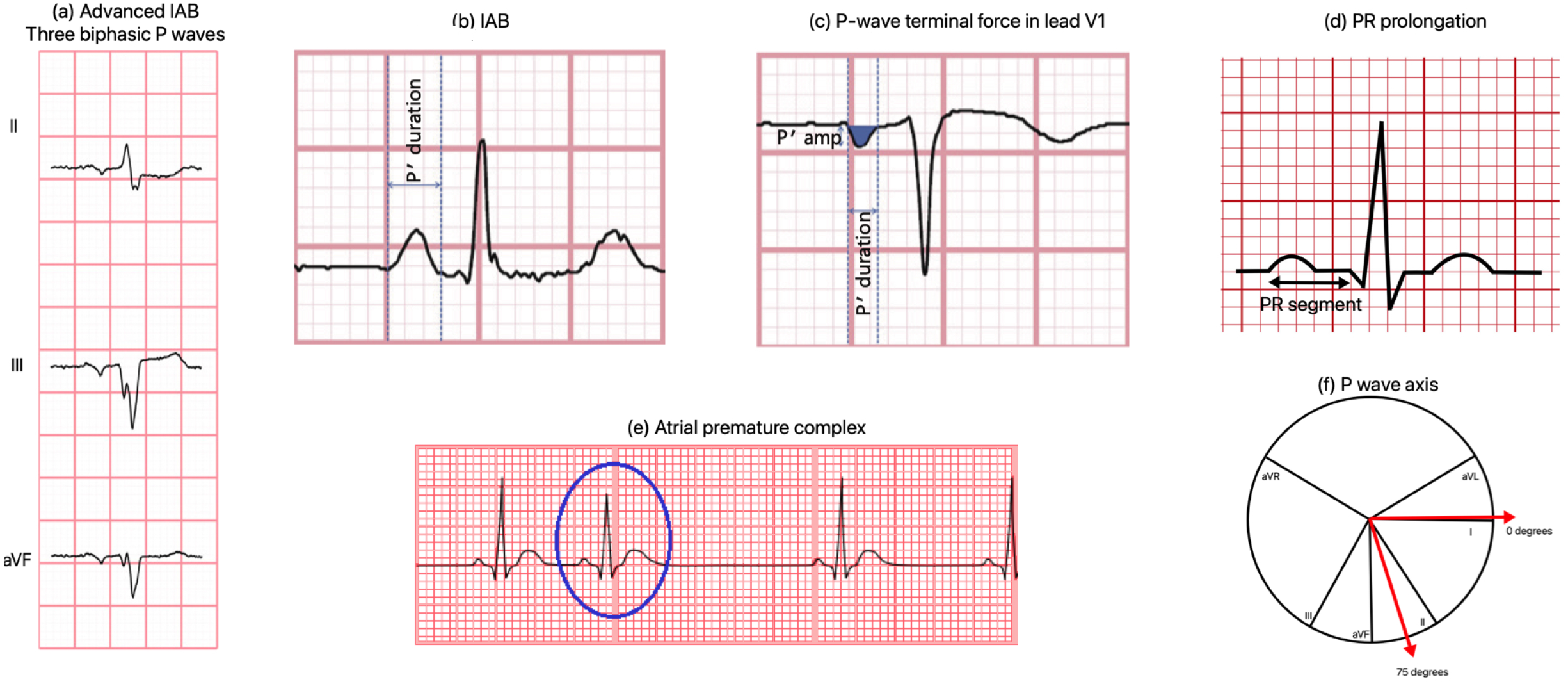
Selected ECG abnormalities. (a) aIAB was defined as P-wave duration > 120ms with biphasic P-waves in leads II, III and aVF. (b) IAB was defined as P-wave duration > 120ms with 1 or more biphasic P-waves. (c) PTFV1 is the area of the negative deflection of the P-wave in lead V1 x duration of this negative deflection (abnormal > 4000 µV × ms) (d) PR prolongation defined as >200ms (e) APC (f) APWA defined as P-wave axis outside 0-75 degrees

### P-wave

#### P-wave terminal force V1 (PTFV1)

Five studies (n= 62,861 patients) identified PTFV1 > 40 mV.ms as a predictor of AF. This included; two cohorts from the community, one cohort of patients with cryptogenic stroke and two ESUS cohorts (14–18). In a meta-analysis of 12 studies, PTFV1 predicted AF over a 14-year follow-up with OR 1.39; 95% CI 1.08-1.79, p=0.01 (19). One study indicated PTFV1 did not predict risk of AF, up to 10-year follow-up period.^52^

#### P-wave duration and Interatrial block (IAB)

Four studies from community cohorts (n=192,308) found advanced interatrial block (aIAB) (defined as P-wave duration ≥120ms with biphasic P-waves in leads II, III and aVF) to be an independent predictor of AF (20–23). This included a metanalysis of 16 studies (n=18 201 patients) with HR 2.58; 95% CI 1.35-4.96, p<0.01 over 15.1 years mean follow up. In this same meta-analysis partial interatrial block (pIAB) was not determined to be a significant predictor of AF.

Six studies (n= 302,257) found pIAB (P-wave duration ≥120ms) or IAB (pIAB pattern with biphasic P-waves in 1-2 of leads II, III and aVF) to be an independent predictor of AF (24–28). One study used P-wave durations of 112 ms and greater to predict AF (26). A further two studies identified pIAB as a predictor of AF in subgroups only; in normal weight individuals(29) (overweight or obese patients were excluded from analysis) and in hypertensive individuals(30).

In three studies, IAB was not significant, though this may have been impacted by small sample sizes (n<300) (15,31). While a meta-analysis found that pIAB did not significantly predict AF, IAB predicted AF with HR 2.42; 1.44-4.07, p=0.001 (23).

#### P-wave dispersion

Two studies (n=585) identified P-wave dispersion as a predictor of AF (28,32,33), while one study in an ESUS cohort did not demonstrate significance on multivariate analysis (15).

#### Abnormal P-wave axis (aPWA)

Four studies (n=28,563) reported abnormal P-wave axis (outside 0-75 degrees) to predict AF. These included two large cohorts drawn from the community, a large cohort of patients with diabetes (n=8965), and one cohort of patients following stroke (n=222) (15,28,34– 36). P-wave axis was not identified as a predictor of AF in one study conducted in an ESUS population. (15)

#### Other P-wave parameters

Other P-wave parameters reported included P-wave amplitude (PWA) (18,32), P-wave area/P-wave duration index (37) and P-wave duration/P-wave vector magnitude (33)

One study reported association with higher PWA and AF, while another reported an association with lower values (18,32). P-wave area/P-wave duration predicted AF at 14.7 years (n=632) with HR 2.80; 95% CI 1.64-4.79, p < 0.001 for development of AF.

#### PR interval prolongation

In seven studies (n=785,927), PR interval prolongation (>200ms) was identified as a significant predictor of developing AF in healthy participants. A further study identified PR interval prolongation as significant but only in the hypertensive subgroup (ref). PR duration was defined as > 200-220 ms(17,18,24,30,38,39). In a large meta-analysis PR interval prolongation was associated with risk of AF with HR 1.3; 95% CI 1-13=1.49, p<0.0001 (40).

#### Ventricular conduction defects (Prolonged QRS and LAFB)

Three studies (n=123,379) found QRS prolongation predictive of incident AF (two in healthy cohorts, one in a cryptogenic stroke cohort). However, secondary analysis of one of these determined that ventricular conduction defects was not significant (41–44). One further study found left anterior fascicular block predictive of AF HR 2.1; 95% CI 1.1-3.9, p=0.023 (45)

#### Left ventricular hypertrophy (LVH)

LVH predicted AF in five studies of healthy patients (17,30,46–48) In one of these studies, LVH was only determined to be significant in an hypertensive population(45).

#### QTc prolongation

Seven studies (n=623,408) reported a significant link between QTc prolongation and risk of AF, including 5 healthy cohorts, one post-stroke cohort and one meta-analysis (30,43,49– 52). In a large meta-analysis, HR at 21 years of follow up was 1.16; 95% CI 1.09-1.24, I2=90%.

#### ST-T segment abnormalities

ST-T segment abnormalities predicted incident AF in four studies of healthy cohorts (30,48,50) with one study finding this not to be significant, also in a healthy cohort of patients (46).

#### Resting bradycardia and high resting heart rate

Resting bradycardia (HR < 50bpm) predicted incident AF in two studies of large healthy cohorts (48,53). In comparison, high resting heart rate predicted incident AF in the same study(53) and in one further study, with cut-off 76 bpm and 72 bpm respectively (53,54).

#### Atrial premature complexes (APCs)

One or more supraventricular extrasystoles on a 10 second 12 lead ECG predicted AF in two studies in ESUS and post stroke cohorts (55,56) and three among healthy cohorts (17,46,48). A meta-analysis did not find significance for APCs (57).

In one study the number of APCs on 12 lead ECG correlated with higher risk of AF in an ESUS cohort (>0-1 APCs HR 1.8; 95% CI 1.06-3.05, >1-2 SVE HR 2.26; 95% CI 1.28-4.01, >2 SVE HR 3.19;95% CI 1.93-5.27 (55).

#### Other ECG abnormalities

Right axis deviation (RAD)(48) and QRS-T wave angle (58) were associated with AF in two separate studies. RAD had a strong association with AF at 3 years (HR 4.74, 1.71-13.12, p<0.01), superseded by extreme right axis deviation (defined as per Minnesota code 2-4) (HR 21.8; 2.97-160, p<0.01)(48).

## Discussion

We conducted a systematic review of ECG patterns that predict the risk of AF in various populations, including healthy populations in primary care settings and patients presenting following stroke. Significant predictors of AF were P-wave terminal force in VI (PTFV1) (HR 5.297; 95% CI 3.249-8.636, p<0.001) and advanced interatrial block (aIAB) (HR 5.014; 95% CI 2.638-9.528, p<0.001). Other consistent predictors of AF were prolonged P-wave duration which reflects interatrial block (IAB), abnormal P-wave axis, PWA, PR prolongation, QRS prolongation, left ventricular hypertrophy, ST-T abnormalities, supraventricular extrasystoles and resting sinus bradycardia.

PTFV1 is defined as the duration from onset of negative deflection in lead V1 to nadir of P-wave (refer Figure 2) and it reflects pressure overload in the left atrium, interatrial conduction delay and left atrial fibrosis. The predictive value of PTFV1 for AF was demonstrated in a meta-analysis (OR 1.39; 95% CI 1.08-1.79, 95% CI 0.01). The role of PTFV1 as a predictor of AF appeared increased if used in conjunction with other ECG markers. For example, combined use of PTFV1 and PWA significantly improved predictive value (HR 11.8; 95% CI 4.88-28.6, p<0.001)(59) while the evidence for PWA independently predicting AF was limited. In addition, the sensitivity of PTFV1 may also be dependent on the patient population assessed. The predictive values of PTFV1 increased in post-stroke patients (OR 1.60; 95% CI 1.14-2.25, p=0.007) and in patients with chronic kidney disease on haemodialysis (OR = 4.89; 95% CI 2.54–9.90, p<0.001)(19).

aIAB was demonstrated to be a significant predictor of AF. This marker reflects fibrosis in the left lower atrium and interatrial septum, which may be a precursor to atrial cardiopathy. The predictive value of other markers of conduction delays between the atrial nodes, undifferentiated IAB or partial IAB (pIAB), is less convincing. The lack of a positive predictive outcome may reflect the small sample size (n<300) in these studies. When the data of these studies were included in meta-analyses, undifferentiated IAB was predictive of AF but pIAB remained insignificant (23).

Similar to PTFV1, the risk factor profile of the study population influences the predictive value of IAB. Age is an independent risk factor for both the aIAB and pIAB (22,29). Increasing age positively correlates with the predictive value for pIAB and aIAB(33). Hypertension increased the predictive value of IAB(30) increasing hazard ratio in one study from non-significant 1.18; 95% CI 0.63–2.20 to 1.43; 95% CI 1.07-1.91, p=0.02. Vascular risk factors also appear to have a cumulative effect with high CHA_2_DS_2_ VASc scores increasing the positive predictive value of IAB (HR 8.11; 95% CI 4.88-13.46, p<0.001)(25). The influence of risk factors is also dependent on the population assessed. IAB was found to be predictive of AF in a sub analysis that excluded obese patients. Another study including only patients with CKD stage IV or V found both aIAB and PTFV1 to not be significant predictors of AF (60).

It is therefore plausible that interatrial block reflects the ECG signature of development of AF as it relates to vascular disease, while other mechanisms of AF (e.g. relating to obesity or right heart strain) do not influence this. These results further suggest that advanced interatrial block is a strong predictor of risk of AF in healthy patients. While partial IAB and undifferentiated IAB may predict AF, this effect is augmented among patients with hypertension and high CHADS VASC.

A U-shaped association is suggested between resting heart rate (RHR) and risk of AF. Three studies reported higher risks of AF at heart rates below 50 bpm and above 72 bpm (48,53,54) among healthy patients. A recent meta-analysis(61) suggested that RHR < 70 corresponded to a relative risk of AF of 1.06 per 10 bpm decrease (RR 1.06; 95% CI 1.03-1.08, p<0.001) while HR < 50 had an increased risk of AF RR 1.16; 95% CI 1.10-1.22, p<0.001. Possible mechanisms included genetic predisposition, autonomic system dysfunction, levels of physical fitness and stroke volume. This may be explained by prolonged vagal activation, which can reduce action potential duration and stabilise re-entrant rotors in the atria, potentiating AF. This supports the correlation between sick sinus syndrome and AF in the elderly (61).

Atrial premature beats may indicate an electrically irritable atrium as sinus rhythm has not suppressed any potential arrhythmogenic potential foci. We identified five studies that demonstrated an increased risk of AF with 1 or more supraventricular ectopic beats (55). This was already identified as a significant predictor of AF on Holter monitoring in a recent meta-analysis(57) (defined as 30 or more atrial ectopic beats per hour, HR 2.96, 95% CI 2.33-3.76).

There have been previous unsuccessful attempts to add ECG parameters to risk prediction scores (such as CHARGE-AF score) (62). Three of the reviewed studies incorporated P-wave abnormalities into a risk prediction score. These demonstrated that PTFV1, P-wave amplitude and aIAB (18,34,59) result in improved predictive value of AF (with PTFV1 and P-wave amplitude employed in all three risk prediction scores). In one study, while the presence of any one of these abnormalities corresponded with a 41% risk of developing AF at 2 years, a second or third abnormality increased this risk to 70%. Other risk prediction scores reviewed which improved the prediction of AF included LVH and supraventricular extrasystoles.(46,48,59) Further refinement of scores incorporating ECG abnormalities may improve detection of AF.

Our study has limitations. Heterogeneity of populations that were included in our study may have accounted for some differences in results. While some studies were strictly post-stroke, other studies recruited patients from primary care settings, with a selection-bias for patients with ECG.

Furthermore, there may have been a misclassification bias, with a lack of standardised follow up of AF across the studies. The majority of studies did not employ invasive cardiac monitoring or have a clearly defined method of assessing for development of AF, which may have resulted in this being underreported.

We identified PTFV1 and aIAB as strong predictors of AF. Other consistent predictors of AF were prolonged P-wave duration, abnormal P-wave axis, P-wave amplitude, PR prolongation, QRS prolongation, left ventricular hypertrophy, ST-T abnormalities, supraventricular extrasystoles and resting sinus bradycardia. While these ECG abnormalities have utility in predicting AF independently, this increases significantly when they are combined with other ECG abnormalities, clinical risk factors (such as age, hypertension or high CHADS-VASC) or both. By using these markers, further development and validation of clinical risk prediction scores may enable us to identify a cohort of patients who present following ESUS in whom we can search aggressively for AF, and make way for prospective trials to determine whether they may benefit from anticoagulation.

## Data Availability

Further data is available from the authors on request.

